# High frequency of SARS-CoV-2 RNAemia and association with severe disease

**DOI:** 10.1101/2020.04.26.20080101

**Authors:** Catherine A. Hogan, Bryan Stevens, Malaya K. Sahoo, ChunHong Huang, Natasha Garamani, Saurabh Gombar, Fiona Yamamoto, Kanagavel Murugesan, Jason Kurzer, James Zehnder, Benjamin A. Pinsky

**Author notes:** **Corresponding author:** Benjamin A. Pinsky, 3375 Hillview, Room 2913, Palo Alto, CA 94304.

## Abstract

**Background:** Detection of SARS-CoV-2 RNA in the blood, also known as RNAemia, has been reported, but its prognostic implications are not well understood. This study aimed to determine the frequency of SARS-CoV-2 RNA in plasma and its association with the clinical severity of COVID-19.

**Methods:** An analytical cross-sectional study was performed in a single-center tertiary care institution in northern California and included consecutive inpatients and outpatients with COVID-19 confirmed by detection of SARS-CoV-2 RNA in nasopharyngeal swab specimens. The prevalence of SARS CoV-2 RNAemia and the strength of its association with clinical severity variables were examined and included the need for transfer to an intensive care unit (ICU), mechanical ventilation and 30-day all-cause mortality.

**Results:** Paired nasopharyngeal and plasma samples were included from 85 patients. The overall median age was 55 years, and individuals with RNAemia were older than those with undetectable SARS-CoV-2 RNA in plasma (63 vs 50 years; p=0.001). Comorbidities were frequent including obesity (37.7%), hypertension (30.6%) and diabetes mellitus (22.4%). RNAemia was detected in a total of 28/85 (32.9%) individual patients, including 22/28 (78.6%) who required hospital admission. RNAemia was detected more frequently in individuals who developed severe disease including the need for ICU transfer (32.1% vs 14.0%; p=0.05), mechanical ventilation (21.4% vs 3.5%; p=0.01) and 30-day all-cause mortality (14.3% vs 0%; p=0.01). No association was detected between RNAemia and estimated levels of viral RNA in the nasopharynx. An additional 121 plasma samples from 28 individuals with RNAemia were assessed longitudinally, and RNA was detected for a maximum duration of 10 days.

**Conclusion:** This study demonstrated a high proportion of SARS-CoV-2 RNAemia, and an association between RNAemia and clinical severity suggesting the potential utility of plasma viral testing as a prognostic indicator for COVID-19.

## Introduction

The coronavirus disease 2019 (COVID-19) is most frequently diagnosed from nasopharyngeal (NP) swab samples. Preliminary data suggest that severe acute respiratory syndrome coronavirus 2 (SARS-CoV-2) RNA may also be detected in plasma and serum.^1,2^ However, the frequency and prognostic implication of viral RNA detection in these sample types is not fully understood. We aim to investigate the association between detectable SARS-CoV-2 RNA in nasopharyngeal swab and plasma samples, and to describe clinical implications of viral RNA detection in plasma.

## Methods

We performed a cross-sectional study at the Stanford Health Care (SHC) Clinical Virology Laboratory, which serves adult and pediatric tertiary care hospitals and affiliated primary care and specialty clinics in northern California. From the beginning of in-house SARS-CoV-2 testing on March 2 2020, residual EDTA whole blood was systematically collected from all SHC patients with a respiratory sample positive for SARS-CoV-2 RNA. NP samples collected with detectable SARS-CoV-2 RNA between March 2 2020 and April 8 2020 were identified and included in the study if residual EDTA whole blood had been collected within 72 hours before or after the NP collection. If available, residual whole blood samples from serial collections, with or without paired NP samples, were also included in the analysis. A volume of 400μL of EDTA plasma from the residual whole blood was extracted and processed to assess for SARS-CoV-2 RNA. Testing was performed based on the adaptation of published rRT-PCR assays targeting the envelope *(E)* gene.^3–5^ The standard cycle threshold (Ct) values of positive tests with this assay range from <20 to 45 cycles. Confirmatory testing of all plasma samples with a Ct value of 40 or greater was performed and resulted as positive if reproducible.^4^ Given that viral culture was not performed to determine the viability of virus, the presence of viral RNA in plasma is referred to as RNAemia instead of viremia. Sample size was based on a convenience set of all available clinical samples without formal power calculation. Statistical analysis was completed using Stata v15.1. Descriptive analysis was performed by Chi-squared test for categorical variables, or Fisher’s exact test when there were less than 5 datapoints per cell, and by Mann-Whitney *U* test for continuous variables. A two-tailed p value of <0.05 was considered significant. This study was approved by the Stanford Institutional Review Board (protocol #48973). Written individual consent was waived for this study.

## Results

A total of 7,240 samples from SHC patients were tested for SARS-CoV-2 corresponding to 4,485 outpatients, 2,452 ED patients, and 303 inpatients, and 453 (6.3%) were positive. Of these, 85 individuals with paired NP and plasma samples were included for analysis (Table 1). The overall median age was 55 years (inter-quartile range (IQR) 40-69) and only two pediatric cases met inclusion criteria, aged 9 and 17 years. Individuals with detectable RNAemia were significantly older than those without RNAemia (63 vs 50 years; p=0.001). Comorbidities were prevalent including obesity in 32/85 (37.6%), hypertension in 26/85 (30.6%) and diabetes mellitus in 19/85 (22.4%); no association was detected between each comorbidity and RNAemia. The overall median number of days of symptoms at the time of NP testing was 7 days (IQR 4-12) and was similar in individuals with and without RNAemia. The most common symptoms were cough (87.1%) and fever (69.8%). RNAemia was detected in a total of 28/85 (32.9%) patients. ICU transfer occurred significantly more frequently in individuals with RNAemia compared to those with undetectable SARS-CoV-2 RNA in plasma (32.1% vs 14.0%; p=0.05). Mechanical ventilation occurred more frequently in individuals with RNAemia (21.4% vs 3.5%; p=0.01), as did mortality (14.3% vs 0%; p=0.01). All 4 deaths occurred in individuals with RNAemia. A total of 15/17 (88.2%) plasma samples with presence of RNAemia in severely ill patients were collected prior to or on the same day as their ICU transfer. For the 16 individuals with suspected community-acquired COVID-19 admitted to the ICU, time from hospital admission to the ICU was short with 13/16 (81.3%) transferred on the same day or within 1 day of admission. No association was detected between estimated viral loads in the nasopharynx, as estimated by RT-PCR Ct value, and detection of RNAemia. Furthermore, plasma *E* gene Ct values were high which is consistent with low viral burden, with a median of 37.5 (IQR 35.1-39.6).

**Table 1.** Characteristics of 85 patients with paired nasopharyngeal and plasma samples tested for SARS-CoV-2

|  |  | **Overall**<br>**patients**<br>**(n=85)** | **Detectable**<br>**plasma RNA**<br>**(n=28)** | **Non-**<br>**detectable**<br>**plasma RNA**<br>**(n=57)** | **p-**<br>**value**<br>**\*** |
| --- | --- | --- | --- | --- | --- |
| Age (median; IQR) |  | 50 (40-69) | 63 (47.5-71) | 50 (37-67) | 0.001 |
| Sex (%) | M | 40 (47.1) | 14 (50.0) | 26 (45.6) | 0.8 |
|  | F | 45 (52.9) | 14 (50.0) | 31 (54.4) |  |
| Location at<br>presentation | ED | 79 (92.9) | 26 (92.9) | 53 (93.0) | 1 |
|  | Outpatient | 5 (5.9) | 2 (7.1) | 3 (5.2) |  |
|  | Inpatient | 1 (1.2) | 0 | 1 (1.8) |  |
| Immunocompromis<br>ed | No | 78 (91.8) | 25 (89.3) | 53 (93.0) | 0.7 |
|  | Yes | 7 (8.2) | 3 (10.7) | 4 (7.0) |  |
| Comorbidities (%<br>present) | Obesity<br>(BMI≥30) | 32 (37.7) | 10 (35.7) | 22 (38.6) | 0.9 |
|  | (Obesity<br>status<br>unknown) | 7 (8.2) | 3 (10.7) | 4 (7.0) |  |
|  | Diabetes<br>mellitus | 19 (22.4) | 7 (25.0) | 12 (21.1) | 0.7 |
|  | HTN | 26 (30.6) | 9 (32.1) | 17 (29.8) | 0.8 |
|  | COPD/asth<br>ma | 12 (14.1) | 3 (10.7) | 9 (15.8) | 0.7 |
| Days of symptoms at time of NP<br>swab collection<br>(median; IQR) |  | 7 (4-12) | 7 (4-14) | 7 (4-11) | 0.6 |
| Symptoms (%<br>present) | Fever | 60 (69.8) | 20 (71.4) | 40 (69.0) | 0.8 |
|  | Cough | 74 (87.1) | 24 (85.7) | 50 (87.7) | 0.8 |
|  | Headache | 21 (24.7) | 4 (14.3) | 17 (29.8) | 0.3 |
|  | (Headache<br>unknown) | 3 (3.5) | 1 (3.6) | 2 (3.5) |  |
|  | Fatigue | 35 (41.2) | 16 (57.1) | 19 (33.3) | 0.1 |
|  | (Fatigue<br>unknown) | 3 (3.5) | 1 (3.6) | 2 (3.5) |  |
|  | GI | 35 (41.2) | 12 (42.9) | 23 (40.4) | 0.8 |
|  | (GI<br>unknown) | 2 (2.4) | 1 (3.6) | 1 (1.8) |  |
| Lymphopenia at<br>presentation | No | 37 (43.5) | 11 (39.3) | 26 (45.6) | 0.6 |
|  | Yes | 48 (56.5) | 17 (60.7) | 31 (54.4) |  |
| Required hospital admission | No | 26 (30.6) | 6 (21.4) | 20 (35.1) | 0.2 |
|  | Yes | 59 (69.4) | 22 (78.6) | 37 (64.9) |  |
| Required ICU transfer | No | 68 (80.0) | 19 (67.9) | 49 (86.0) | 0.05 |
|  | Yes | 17 (20.0) | 9 (32.1) | 8 (14.0) |  |
| Required mechanical ventilation | No | 77 (90.6) | 22 (78.6) | 55 (96.5) | 0.01 |
|  | Yes | 8 (9.4) | 6 (21.4) | 2 (3.5) |  |
| 30-day all-cause mortality (%) | No | 81 (95.3) | 24 (85.7) | 57 (100) | 0.01 |
|  | Yes | 4 (4.7) | 4 (14.3) | 0 |  |
| Nasopharyngeal CT value | CT <25 | 24 (28.2) | 11 (39.3) | 13 (22.8) | 0.2 |
|  | CT 25-35 | 36 (42.4) | 12 (42.9) | 24 (42.1) |  |
|  | CT >35 | 25 (29.4) | 5 (17.9) | 20 (35.1) |  |
| Median NP CT (IQR) |  | 30.0 (23.9-35.7) | 27.1 (21.1-34.4) | 30.2 (25.8-36.2) | 0.1 |
| Median plasma E gene CT (IQR) |  | - | 37.5 (35.1-39.6) | - | - |

\*Chi-squared, Fisher's exact test or Mann-Whitney *U* test
COPD: chronic obstructive pulmonary disease; Ct: cycle threshold; CVD: cerebrovascular disease;
DM: diabetes mellitus; *E* gene: envelope gene; F: female; HTN: hypertension; ICU: intensive care
unit; IQR: interquartile range; M: male; mL: milliliter; *N* gene: nucleocapsid gene; NP:
nasopharyngeal; RNA: ribonucleic acid.

An additional 121 plasma samples from the 28 individuals with RNAemia detected on the paired plasma sample were tested to assess longitudinal trends. Each individual underwent a median of 2 (IQR 0-7) repeat SARS-CoV-2 plasma tests, with a range of 1 to 27 days between tests. The longest period of RNAemia was 10 days and occurred in an ICU-admitted patient. SARS-CoV-2 RT-PCR Ct values were comparable to those seen in the initial paired samples with a median of 37.2 (IQR 34.6-39.6). Detailed paired NP and plasma values are presented in Figure 1 for the individuals with 5 or more longitudinal samples including one sample collected prior to ICU transfer.

**Figure 1.**
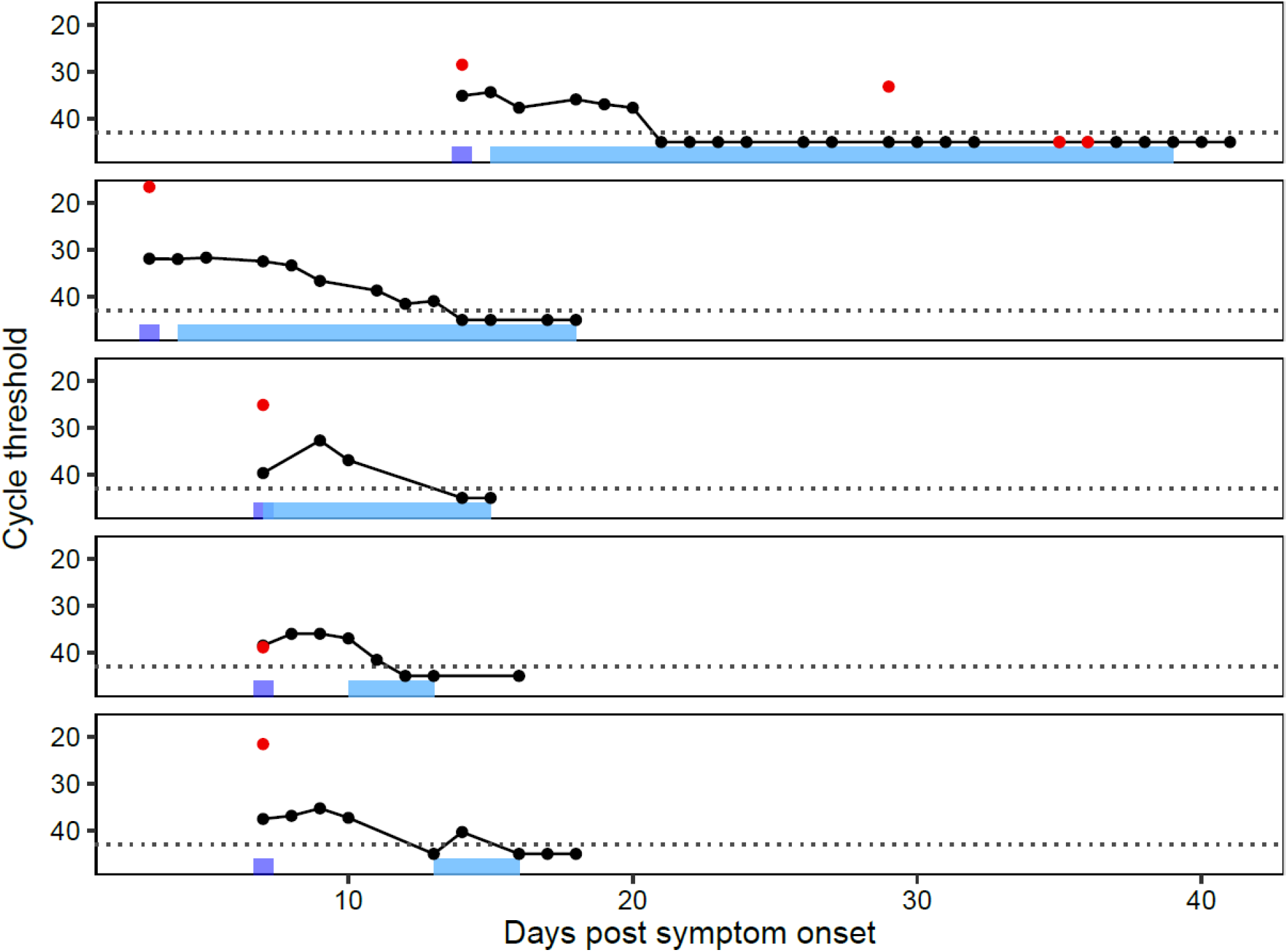
Longitudinal SARS-CoV-2 nasopharyngeal and plasma data for individuals with 5 or 213 more samples including at least one sample collected prior to ICU transfer The dotted background line represents the real-time reverse transcriptase polymerase chain reaction threshold. Black circles represent the cycle threshold result of plasma samples tested for SARS-CoV-2. Red dots represent the cycle threshold result of nasopharyngeal samples tested for SARS-CoV-2. The purple rectangles represent the time of hospital admission. The light blue rectangles represent the timeline of ICU stay.

## Discussion

This analytical cross-sectional study assessed 85 individual with a nasopharyngeal swab positive for SARS-CoV-2 RNA for detection of RNAemia in paired plasma samples. Thse findings show that RNAemia was present in almost one third of samples tested. This proportion is higher than previously reported from a cohort of COVID-19-positive individuals in China, where only 3/307 (1%) of plasma samples tested for SARS-CoV-2 were detected.^1^ Other small studies have reported varying rates of RNAemia from plasma (0-20%), whole blood (8-40%) and serum samples (0-17%).^2,6–11^ However, direct comparison of these results is challenging due to the use of different testing protocols, including differences in specimen extraction volumes (ranging from 80 to 200μL), sample types examined (plasma, serum and whole blood), viral genomic targets tested (N, *S, E* or *ORF1ab* genes), variable timing of blood collection relative to symptom chronology and different patient populations. For example, using the most similar plasma RT-PCR method to this study as a comparator, the higher prevalence of RNAemia may be explained by the higher proportion of individuals with severe disease in this cohort (ICU transfer 20.0% vs 11.1%).^8^ By comparison, clinical illness with SARS-CoV-1 was almost always severe, and RNAemia was reported to occur in up to 75% of plasma samples between 5 and 7 days of illness, and up to 78% of serum samples on admission.^12–14^

The association between RNAemia, ICU transfer and mechanical ventilation observed in this study suggests the potential utility of plasma SARS-CoV-2 RNA testing as a prognostic indicator. Indeed, although this study was not performed in a controlled setting, most (88.2%) plasma samples with presence of RNAemia in severely ill patients were collected prior to or on the same day as their ICU transfer, supporting a possible role for the early detection of at-risk individuals. Preliminary data from a limited number of patients have shown an association between RNAemia and severe COVID-19, and support the presence of RNA in extra-pulmonary sites.^11^ A similar association between detection of SARS-CoV-1 RNA in serum and clinical complications including oxygen desaturation, mechanical ventilation and mortality was noted in 2004.^15^ Thus, plasma may be considered as a complementary modality for the early identification of individuals likely to develop severe COVID-19.

This study is limited by its single-center experience, and use of a convenience set of samples for SARS-CoV-2 testing that favored overall selection of individuals with more severe disease given the need for having had a blood sample collection. However, that a signal for ICU transfer was detected despite this patient selection lends further support to the clinical importance of RNAemia. Furthermore, individuals in this cohort rapidly required ICU transfer from their time of arrival to the hospital, such that plasma samples collected a single day before or on the same day as ICU transfer reflect real-world experience. In addition, in the absence of viral culture data, it cannot be determined at present whether detectable RNA represents intact infectious virus or inactive, non-replicating nucleic acid. A recent study attempted to address this question but failed to detect SARS-CoV-2 RT-PCR in all serum samples such that culture could not be pursued.^9^ Further work should thus be performed to investigate this question given the potential important biosafety and blood product transfusion implications, though non-severely ill individuals are less likely to be RNAemic.^16^

In summary, this study demonstrated a high overall proportion of detectable SARS-CoV-2 RNAemia, and an association between plasma viral detection and older age, and severe clinical disease including need for ICU transfer, mechanical ventilation and 30-day all-cause mortality. Further studies are required to investigate the prognostic potential of RNAemia to facilitate early therapeutic and supportive interventions prior to the development of severe clinical manifestations of COVID-19.

## Data Availability

The data generated and analyzed during the current study are available from the corresponding author on reasonable request.

